# IVF-Conceived Offspring Exhibit Altered LINE-1 Retrotransposition Dynamics Associated with Long-Term Disease Risks

**DOI:** 10.1101/2025.10.21.25338362

**Authors:** Jian Li, Jia Huang, Zhenglin Zhang, Jiefeng Xian, Ying Liu, Qingxue Zhang, Hui Chen, Yabin Guo, Yu Li

## Abstract

**Background:** In vitro fertilization (IVF) has revolutionized reproductive medicine; however, IVF-conceived offspring exhibit a higher risk of various long-term health issues. The underlying mechanisms remain unclear. Long Interspersed Nuclear Element-1 (L1), a mobile genetic element sensitive to environmental stress, is a potential mediator. We hypothesized that the IVF procedure acts as an embryonic stress, altering L1 retrotransposition and contributing to genomic instability associated with disease risk.

**Methods:** Whole blood from 33 IVF and 42 naturally conceived (NC) offspring were collected for deep sequencing. We quantified L1 genomic content and mapped novel L1 insertions and deletions with Bowtie2 and MELT, and further compared their frequencies and genomic distributions between the two groups. Gene-disease association analysis was performed on genes within 500kb of differential L1 sites.

**Results:** The overall L1 content was significantly higher in IVF offspring compared to NC controls (P<0.05), a finding robustly confirmed in three sibling pairs from the same parents where the IVF children had higher L1 content than their NC siblings. We identified 11 specific genomic loci with significantly different L1 insertion frequencies and 14 loci with different deletion frequencies between IVF and NC offspring. Notably, these differential sites were often located near genes significantly associated with metabolic, cardiovascular, neuropsychiatric, and neoplastic diseases, conditions previously linked to IVF conception.

**Conclusion:** Our findings demonstrate that IVF conception is associated with increased L1 content and altered genomic distribution of L1 elements in offspring. The disease-related genes near these aberrant L1 sites provide a plausible genomic link between IVF-associated embryonic stress and the increased risk of specific diseases in the IVF population. This study offers novel insights into the molecular safety of ART.

## Introduction

Currently, more than 15% of couples suffer from infertility or subfertility^1^. Over 8 million babies have been born through assisted reproductive technologies (ART) since the invention of in vitro fertilization (IVF) by Robert Edwards and Patrick Steptoe in 1978^2,3^. An increasing number of infertile couples have successfully conceived and delivered offspring through IVF with the development and widespread application. However, alongside these successes, concerns have emerged regarding potential long-term health consequences for the offspring. It has been reported that offspring conceived through IVF have a significantly higher risk of miscarriage, preterm birth, intrauterine growth restriction, low birth weight, and perinatal mortality during pregnancy and the perinatal period compared to those conceived naturally^4–6^. Moreover, IVF-conceived offspring have an increased risk of developing neurological and psychiatric disorders, cardiovascular diseases, asthma, autism, diabetes, obesity, and tumors during their growth and development^7–13^. Therefore, exploring whether IVF is the cause for the higher incidence of these diseases in IVF offspring is of great significance for clarifying the safety of IVF for human offspring and guiding the development of IVF technology in a safer direction.

Recent studies have found that transposable elements, the “dark matter” in the genome, play an important role in biological activities^14,15^. Among them, Long Interspersed Nuclear Element-1 (LINE-1 or L1) has garnered significant research interest. The L1 gene is approximately 6,000 base pairs (bp) in length, and there are around 500,000 copies in the human genome, accounting for about 17% of the entire genomic sequence. L1 is the most abundant transposable element in the human genome and the only active autonomous transposable element^16,17^. L1 transposition mainly occurs during embryonic development, generating new integration sites in the genome and resulting in different L1 integration sites in somatic cells^18–20^. L1 transposition was reported to be more active in the hippocampus region of mouse pups that lacked maternal care in 2018^21^. This suggests that early-life stress can increase L1 transposition, and stress-induced activation of transposable elements is a relatively common phenomenon^22^. Considering that IVF also represents a significant environmental stress for early embryos, with *in vitro* culture conditions and media composition being markedly different from those *in vivo*, and temperature fluctuations as well as transplantation procedures also being stressful for embryos, we wondered whether the stress associated with IVF activates L1 transposition, leading to more L1 insertions in IVF offspring compared to naturally conceived offspring. It has also been reported that the distribution and activation of L1 are associated with neuronal activity, neuropsychiatric disorders, cardiovascular diseases, metabolic diseases, and tumors^23–28^. Given that IVF procedures subject preimplantation embryos to various non-physiological conditions, including specific culture media, gas concentrations, and temperature fluctuations, we postulated that IVF could serve as a potent activator of L1 retrotransposition. To test this hypothesis, we collected whole blood from offspring conceived through IVF and naturally, then performed deep whole-genome DNA sequencing to detect and compare the content and distribution of L1 in them. We aim to explore whether there is a higher proportion of L1 in the genomes of IVF individuals and whether there are differences in L1 distribution between IVF and naturally conceived populations, which may be a potential source of the increased incidence of various long-term diseases in the IVF offspring population.

## Materials and Methods

### 1. Study population

Umbilical cord blood or peripheral blood were collected from 33 IVF-conceived offspring who received assisted reproductive technology at the Reproductive Center of Sun Yat-sen Memorial Hospital, Sun Yat-sen University, and 42 naturally conceived (NC) offspring delivered successfully at full term (≥37 weeks) in the Department of Obstetrics of Sun Yat-sen Memorial Hospital, Sun Yat-sen University, and Yunfu People’s Hospital from January 1, 2015, to December 31, 2021. Please find the basic information in Table 1. The study was approved by the Research Ethics Board at Sun Yat-sen University affiliated Sun Yat-sen Memorial Hospital. Written informed consent was obtained from their parents.

**Table 1.**
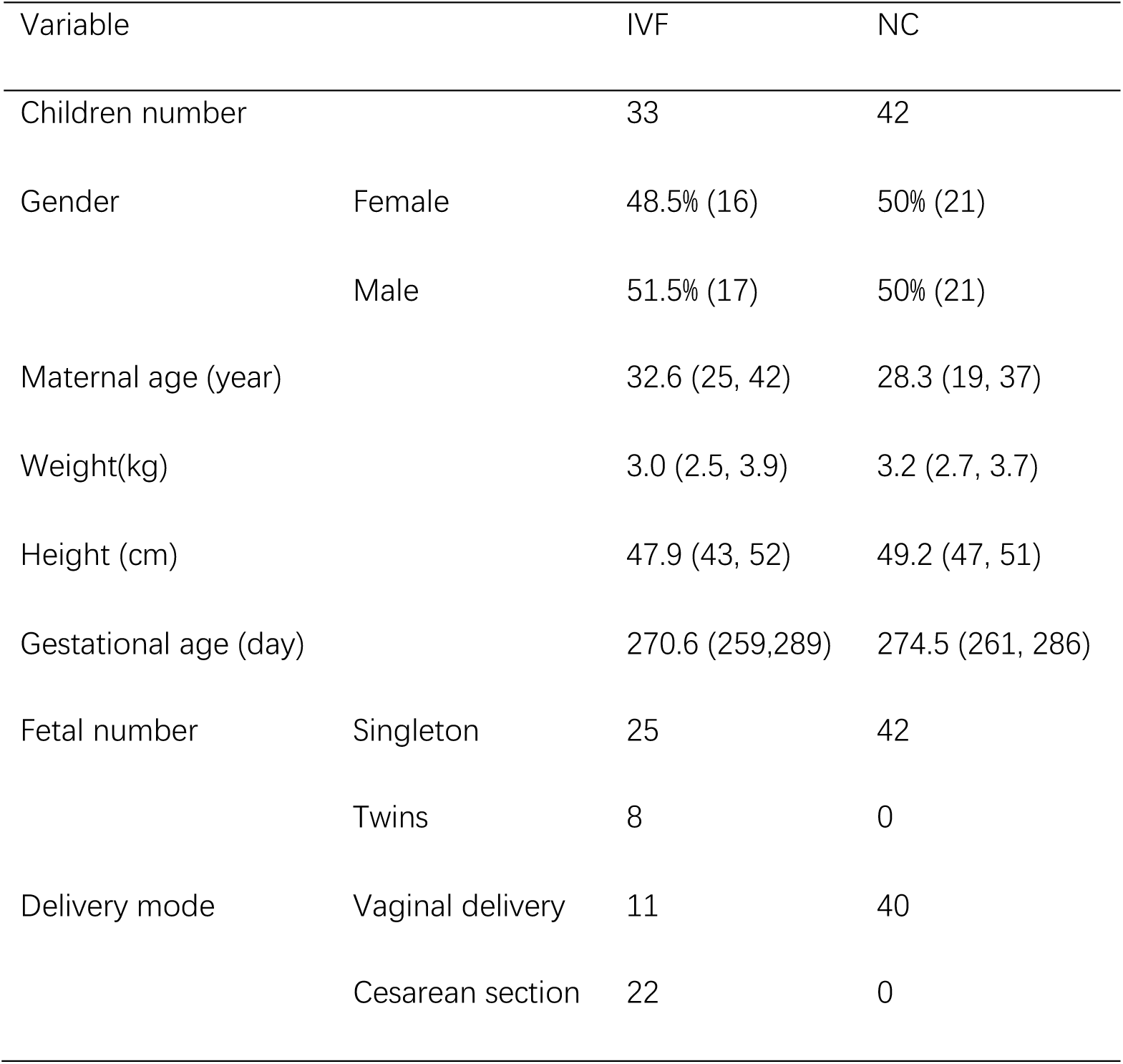
Basic information of the population.

| Variable |  | IVF | NC |
| --- | --- | --- | --- |
| Children number |  | 33 | 42 |
| Gender | Female | 48.5% (16) | 50% (21) |
|  | Male | 51.5% (17) | 50% (21) |
| Maternal age (year) |  | 32.6 (25, 42) | 28.3 (19, 37) |
| Weight(kg) |  | 3.0 (2.5, 3.9) | 3.2 (2.7, 3.7) |
| Height (cm) |  | 47.9 (43, 52) | 49.2 (47, 51) |
| Gestational age (day) |  | 270.6 (259,289) | 274.5 (261, 286) |
| Fetal number | Singleton | 25 | 42 |
|  | Twins | 8 | 0 |
| Delivery mode | Vaginal delivery | 11 | 40 |
|  | Cesarean section | 22 | 0 |

### 2. Whole Genome Sequencing (WGS)

Genomic DNA was extracted from 250 μL umbilical cord blood or peripheral blood using the method and reagents provided by the Whole Blood Genomic DNA Extraction Kit (Solarbio, China). The high-quality purified genomic DNA was then subjected to 10x WGS on the Illumina platform (PE150) by Novogene Bioinformatics Technology Co., Ltd. in Beijing. The clean sequencing data obtained in the form of FASTQ files were used for further analysis.

### 3. L1 detection

To detect L1 proportion in individual genome, we used the full-length L1 sequences downloaded from l1base.charite.de and the L1 sequences from the human reference genome hg38 obtained from the UCSC Genome Browser (https://genome.ucsc.edu/) to build index. We then employed the Bowtie2 software (version 2.2.5, https://bowtie-bio.sourceforge.net/bowtie2/index.shtml) to detect L1 proportion with the command of : “bowtie2 --local -p 32 -x /mnt/d/index -U /mnt/d/XX.fq.gz -S /mnt/d/XX.sam” ^29(p2)^.

### 4. Detect new L1 insertions

We used the hg38 reference sequence provided by MELT to detect new L1 insertions in each sample. Briefly, we first aligned the clean FASTQ files to the Hg38 reference genome in paired-end mode using the BWA software to generate SAM files. Then, we converted the SAM files to BAM files using the samtools software (https://github.com/samtools/samtools)^30^. Finally, we used the LINE1_MELT reference sequence provided by MELT as the index to identify new L1 insertions^31^.

### 5. Detect L1 deletion

After generating BAM files from the clean FASTQ files using the aforementioned method, we employed the MELT-Deletion method provided by MELT, along with its reference L1 sites, to detect L1 deletion sites in the IVF and NC offspring population^32,33^.

### 6. Gene function analysis

After identifying the genes within 500-kilobase (kb) upstream and downstream of the aforementioned sites in the Hg38 genome, we used the GAD_DISEASE_CLASS tool in DAVID to perform gene-disease association analysis and identify potential diseases associated with these genes^34,35^.

### 7. Statistical analysis

Statistical analysis was performed using the R language (version 4.1.2). For continuous variables in normal distribution, comparisons between two groups were analyzed with the independent t-test. For continuous variables not in normal distribution, the Mann-Whitney U test (rank-sum test) was performed. Paired samples were analyzed with the paired t-test (Pair-T Test). Categorical data were presented as percentages and counts, and comparisons between two groups were performed with the Chi-squared test or Fisher’s Exact Test, as appropriate. A p-value of less than 0.05 was considered statistically significant.

## Results

### 1. L1 proportion in IVF offspring genome is higher than that in NC offspring

In this study, we collected a total of 33 umbilical cord blood or peripheral blood from IVF-conceived offspring and 42 samples from naturally conceived offspring. Through whole genome sequencing and alignment with the L1 sequences, we found that L1 proportion in the IVF offspring genome was significantly higher than that in the naturally conceived population (P<0.05, T test) (Fig 1A). Since L1 proportion on the X chromosome is much higher than that on autosomes, we further compared L1 proportion in different genders of IVF and NC populations. We found that L1 proportion in female IVF population was significantly higher than that in female NC population (P<0.05, T test) (Fig 1B), while L1 proportion in male IVF population was slightly higher than that in male NC population but did not reach statistical significance (Fig 1C). There were three pairs of IVF and NC offspring from the same parents among them. The pairwise comparison results demonstrated that L1 proportion in IVF offspring was higher than that in their NC siblings from the same parents (Fig 1D). Moreover, in all three pairs, the IVF offspring were born earlier than the NC offspring, indicating that the higher L1 proportion in IVF offspring was not due to parental age.

**Fig. 1.**
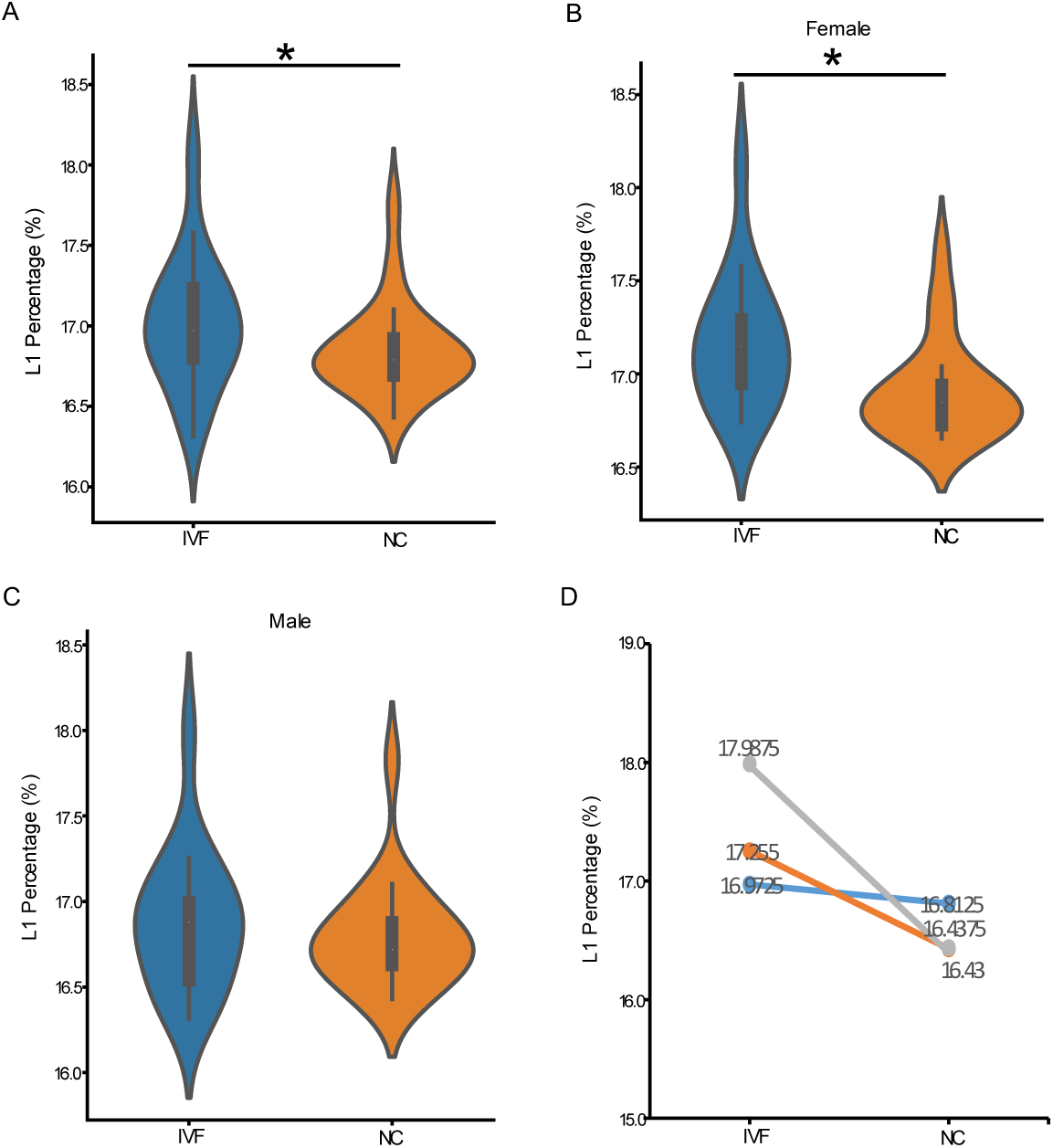
Proportion of L1 in IVF and NC Population. **A,** Proportion of L1 in IVF population is higher than that in NC population (*, P<0.05). **B,** Proportion of L1 in female IVF population is higher than that in female NC population (*, P<0.05). **C,** Proportion of L1 in male IVF population and male NC population. **D,** L1 proportion in three IVF individuals are higher than their naturally conceived siblings the same parents.

### 2. L1 possesses different new insertion sites in IVF and NC offspring

It has been reported that L1 can autonomously transpose into certain gene loci, thereby affecting the function of the corresponding genes^36,37^. After detecting different L1 contents in IVF and NC populations, we further explored whether the distribution of L1 is consistent in these two populations. We used MELT to detect L1 new insertions in all samples. We found that comparing with the reference genome, L1 generated a total of 560 new insertion sites on autosomes and ChrX in these 75 samples (Fig 2A). 322 (57.5%) new insertion sites were located in non-genic regions, and 238 (42.5%) were located in regions encoding genes or producing miRNA, LINCRNA, and antisense RNA. The number of new insertion sites on each chromosome and their distribution patterns are shown in Fig 2A. L1 can be divided into different subfamilies based on the similarity of sequence homology and the order of appearance. We further explored the subfamily affiliation of these new insertion L1. We found that 50% of new insertion L1 belonged to L1Ambig, which could not be accurately classified into a specific L1 subfamily, 35% belonged to the evolutionary younger and more active L1Ta subfamily, and 15% belonged to the even younger and more active L1Ta1d subfamily (Fig 2B). The insertion location of these new L1Ambig, L1Ta, and L1Ta1d members in the genome were also different: about 60.6% of L1Ambig members were inserted into non-genic regions, and 49.4% of L1Ta1d members were inserted into non-genic regions. Even in genic regions, the insertion site preference of each subfamily member was also different. For example, the proportion of L1Ambig members inserted into intron was 33% of all insertions, while the proportion of L1Ta1d members inserted into intron reached 44.7% (Fig 2C-2E). It indicates that the younger and more active L1Ta1d subfamily tends to insert into genic regions. This preference for genic regions by evolutionary younger, active subfamilies suggests a greater potential for IVF-induced L1 insertions to directly disrupt gene function.

**Fig. 2.**
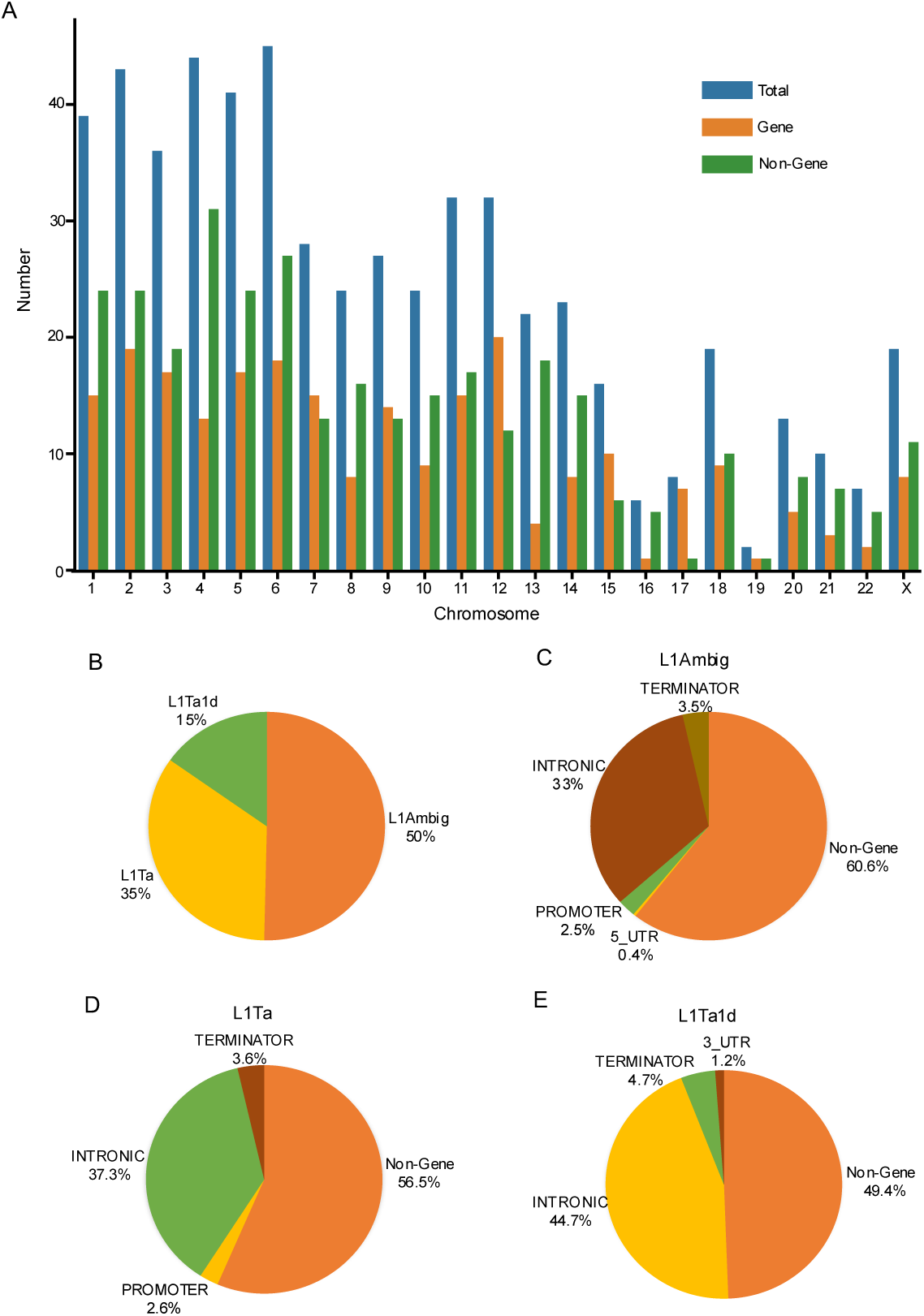
Scheme of L1 new insertion in the studied population. **A,** Distribution of L1 new insertion in different chromosomes in the whole investigation population. **B,** Proportion of L1 subfamily in the whole new insertion sites. **C,** New inserted L1Ambig distribution in genomic location. **D,** New inserted L1Ta distribution in genomic location. **E,** New inserted L1Ta1d distribution in genomic location.

Further exploration revealed that the insertion frequency of L1 at new sites varies among individuals. For instance, insertions were found in all individuals at sites such as chr1_81390097 on Chr1 and chr3_188117521 on Chr3, while insertions were only detected in one individual at sites like chr1_50169914 on Chr1 and chr2_178321473 on Chr2. We further investigated the insertion frequency of each site in the IVF and NC populations (Fig 3). Results revealed that there were significant differences in insertion frequency at 11 sites between the IVF and NC populations (P<0.05) (Fig 4A). Six sites, such as chr8_118709464 on Chr8 and chrX_18334634 on ChrX, *etc*, demonstrated higher insertion frequency in IVF offspring than that in the NC population. Conversely, the insertion frequency in the NC population was significantly higher than that in the IVF population at the other five sites, including chr15_47215145 on Chr15 and chr4_84396322 on Chr4, *etc*.

**Fig. 3.**
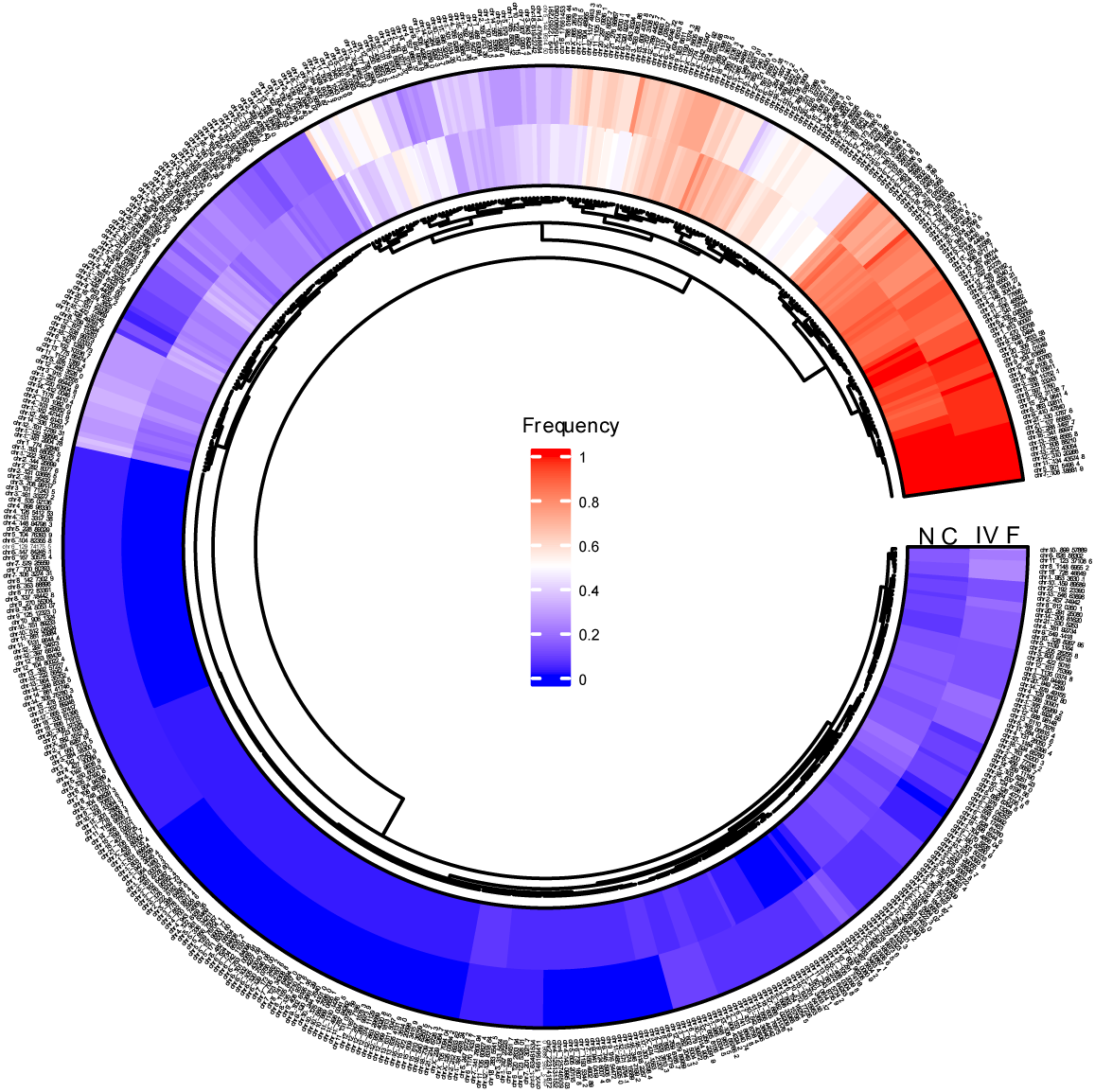
L1 insertion frequency in IVF or NC population in detailed sites.

**Fig. 4.**
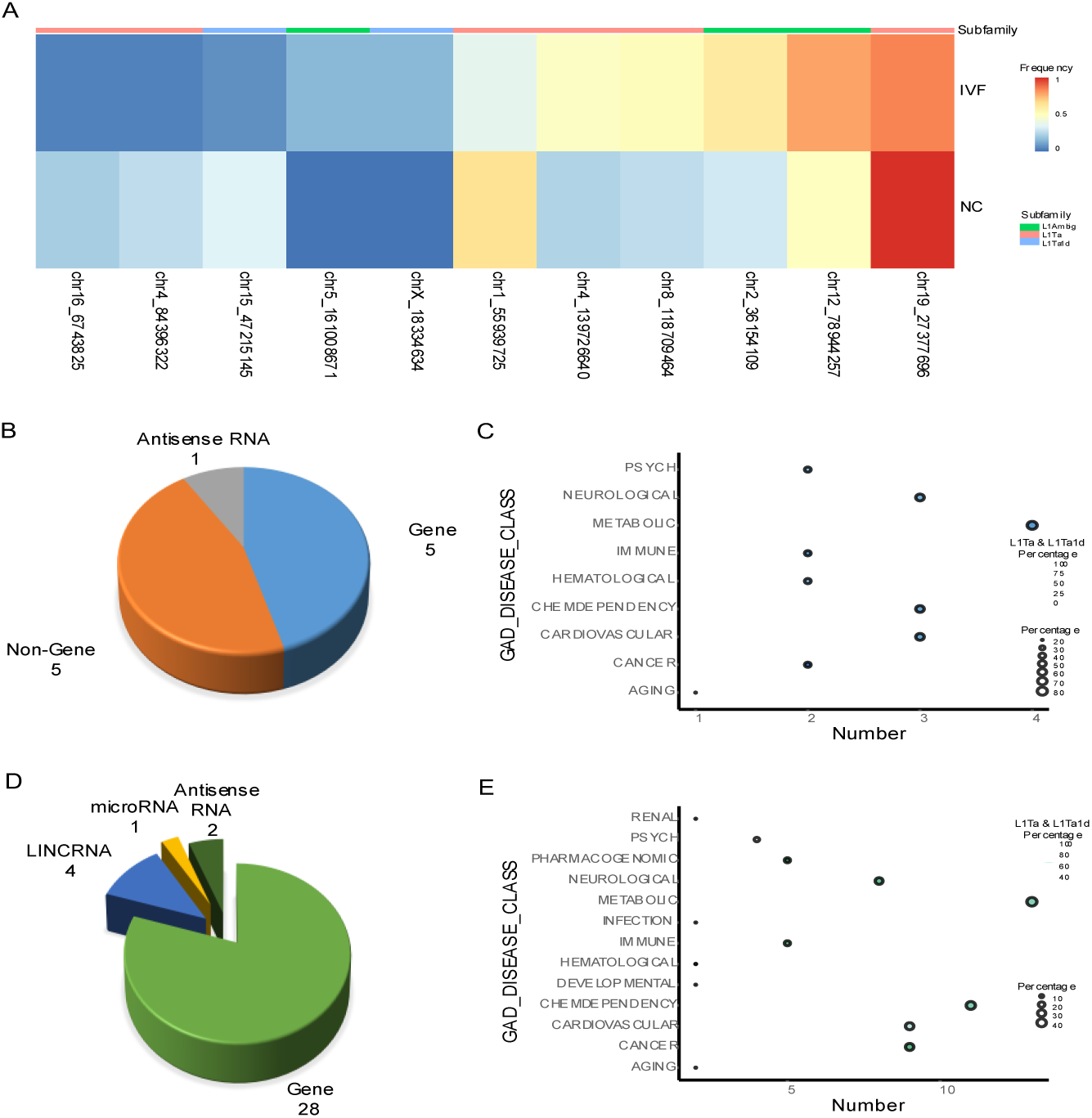
Different insertion of L1 in IVF and NC population. **A,** L1 insertion frequency significantly different in IVF and NC population in 11 sites (P<0.05). **B,** Genomic information of the 11 sites. **C,** Major diseases related to the insertion site genes. **D,** Genomic information around 500kb of the 11 insertion sites. **E,** Major diseases related to genes around 500kb of the 11 insertion sites.

### 3. The functions of genes near different L1 insertion sites in IVF and NC offspring are related to prevalent diseases in IVF offspring

Clinical investigations have revealed that the incidence of metabolic diseases, cardiovascular diseases, and tumors is higher in the IVF offspring population than that in the naturally conceived population. L1 insertions can regulate the expression and function of genes at the insertion sites in various ways, thereby affecting the body’s vital activities. We analyzed the characteristics of these different insertion sites and found that among the 11 sites, 5 sites are in genic regions, and 1 site is in the coding region of SAMD12 antisense RNA 1 (SAMD12-AS1). SAMD12-AS1 has been reported to be associated with the occurrence of liver cancer and neuroblastoma^38,39^. Through gene-disease association analysis using GAD_DISEASE_CLASS in DAVID, we found that among the 5 genes at these sites, 4 are associated with metabolic diseases, 3 with cardiovascular diseases, 3 with neuropsychiatric diseases, and 2 with hematologic diseases, *etc.* (Fig 4C).

Recent research has shown that L1 not only affects the function of genes at the insertion sites but also influences the expression of genes within 500-kb region^40^. Therefore, we further searched and explored genes, antisense RNA, microRNA, and LINCRNA within 500 kb upstream and downstream of these sites. We found that there was a total of 28 genes, 2 antisense RNA, 1 microRNA, and 4 LINCRNA in this region (Fig 4D). Among these 28 genes, 13 are associated with metabolic diseases, 9 with cardiovascular diseases, 9 with tumors, 8 with neuropsychiatric diseases, 2 with development, *etc.* (Fig 4E). The significant enrichment of genes near differential L1 sites in pathways related to metabolism, cardiovascular function, and neurodevelopment provides a plausible genomic link between the observed L1 alterations and the diseases prevalent in the IVF population.

### 4. L1 has different deletion sites in IVF and NC offspring

Furthermore, we used MELT-Deletion to detect the deletion of L1 in the two populations. Compared with the reference genome, we found that there were 460 L1 sites with different degrees of deletion on the autosomes and ChrX in these 75 samples. Fig 5 shows the different deletion frequencies of IVF and NC population at each deletion site. There were significant differences in deletion frequency at 14 sites between the IVF and NC population (P<0.05) (Fig 6A). At four sites, such as chr1_247687172, there was no deletion in the IVF population, while there was partial deletion in the NC population. At two sites, there were deletions of different degrees in both populations, but the deletion frequency was higher in the NC population. At the chrX_78441360 site, the deletion frequency in the IVF population was higher than that in the NC population. At seven sites, such as chr1_247687163, there were deletions of different degrees in the IVF population, while there was no deletion in the NC population.

**Fig. 5.**
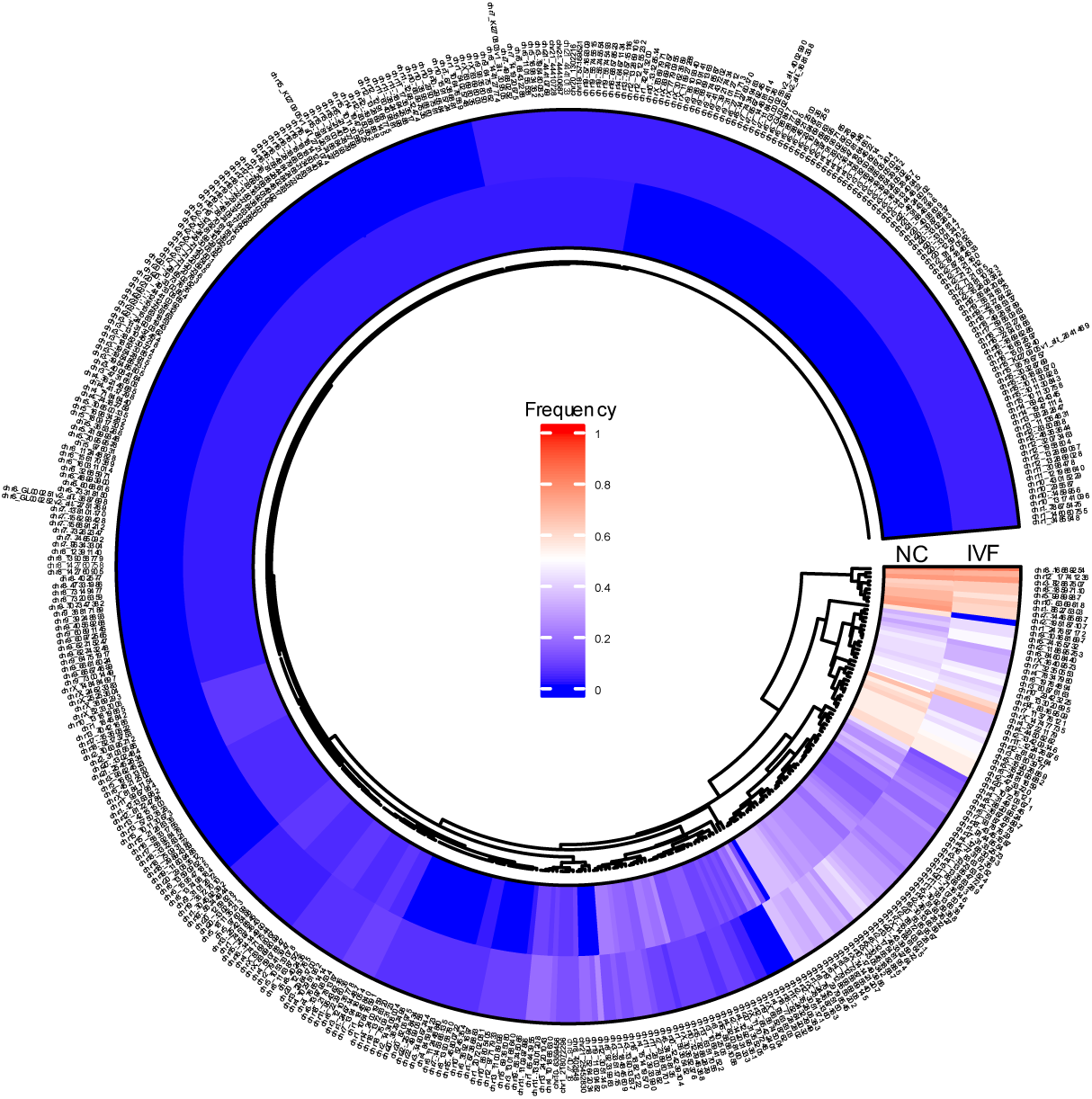
L1 deletion frequency in IVF or NC population in detailed sites.

**Fig. 6.**
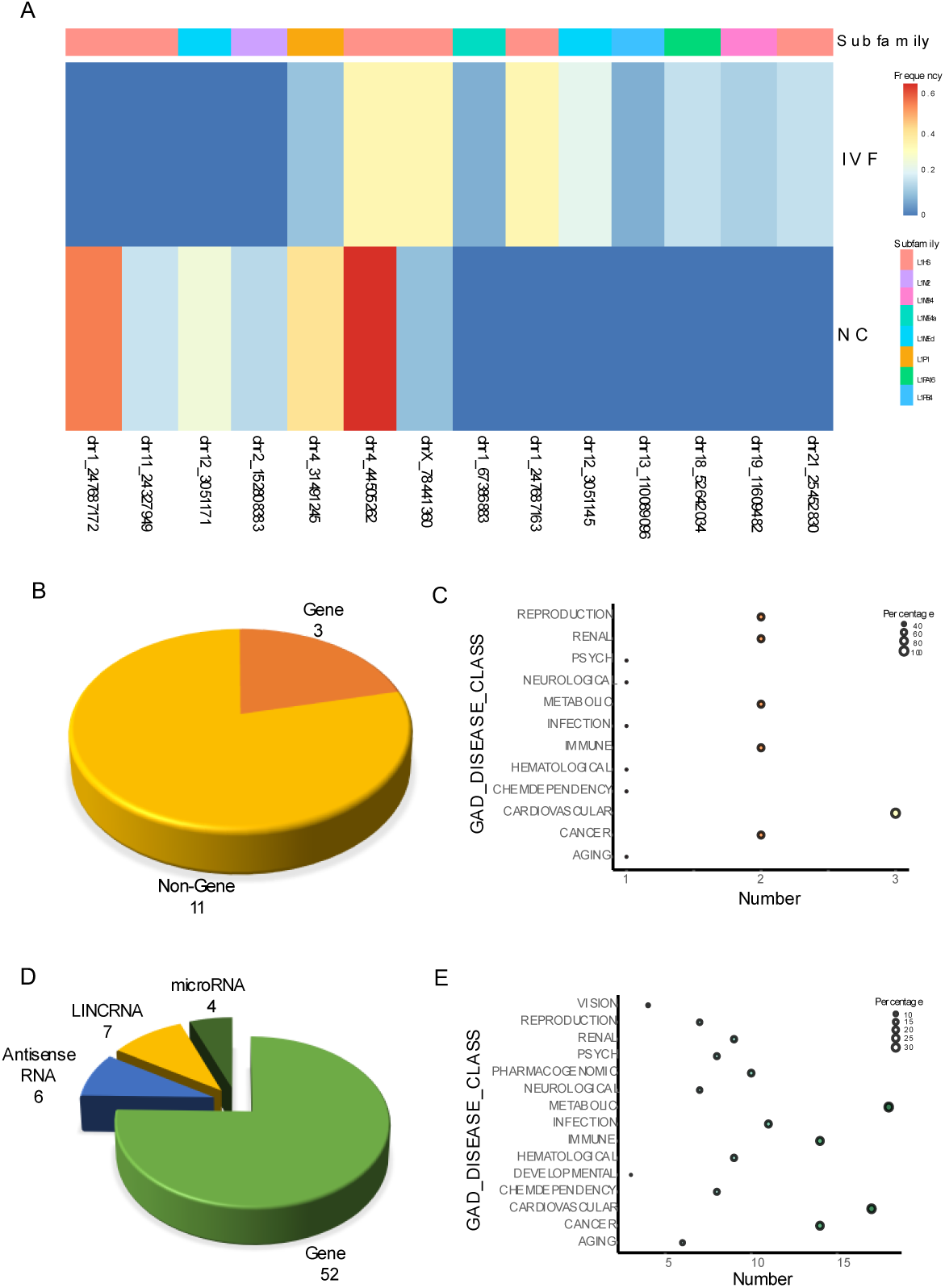
Different deletion of L1 in IVF and NC population. **A,** L1 deletion frequency was significantly different between IVF and NC population in 14 sites (P<0.05). **B,** Genomic information of the 14 sites. **C,** Major diseases related to the deletion site genes. **D,** Genomic information around 500kb of the 14 sites. **E,** Major diseases related to genes around 500kb of the 14 sites.

### 5. The functions of genes near different L1 deletion sites in IVF and NC offspring

Not only does the insertion of L1 affect the expression of its site and nearby genes, L1 deletion also impacts the expression and function of them^41^. Therefore, we further analyzed and explored the genes and their related functions within 500-kb region of these different deletion sites. Among the 14 sites with different deletion frequencies between the IVF and NC population, 3 sites are in genic regions, and 11 sites are in non-genic regions (Fig 6B). According to gene-disease association analysis in DAVID, the functions of the 3 genes at the genic sites are all associated with cardiovascular diseases, 2 with metabolic diseases, and 2 with tumors, *etc.* (Fig 6C). We further searched and explored the information on genes, antisense RNA, microRNA, and LINCRNA within 500 kb upstream and downstream of these 14 different L1 deletion sites. The results demonstrated that there are a total of 52 genes, 6 antisense RNA, 4 microRNA, and 7 LINCRNA in this region (Fig 6D). Among these 52 genes, 18 are associated with metabolic diseases, 14 with tumors, 17 with cardiovascular diseases, 10 with neuropsychiatric diseases, etc. (Fig 6E).

## Discussions

Since the inception of IVF, over 8 million babies have been born, yet concerns persist regarding the potential increased health risks for these offspring. Epidemiological studies have indicated a higher incidence of various adverse perinatal outcomes and long-term diseases, such as metabolic, cardiovascular, and neuropsychiatric disorders, in the IVF-conceived population compared to their naturally conceived counterparts. While the exact mechanisms remain unclear, the unique in vitro environment, characterized by suboptimal culture conditions, fluctuations in temperature, and exposure to light and atmospheric oxygen, is considered a significant environmental stress during the critical window of early embryonic development. This stress may induce epigenetic alterations or genomic instability, contributing to the observed phenotype differences. Given that early-life stress has been shown to activate transposable elements like L1, we hypothesized that the IVF procedure itself might act as a stress, leading to aberrant L1 activity, which could be a potential molecular link to the increased disease risks. In this study, we provide evidence supporting this hypothesis. We found that the overall L1 content in the genomes of IVF offspring was significantly higher than in naturally conceived offspring. This finding was further strengthened by the analysis of three sibling pairs from the same parents, where the IVF-conceived children consistently exhibited a higher L1 proportion than their later-born, naturally conceived siblings. This within-family comparison effectively controls for genetic background and, crucially, rules out confounding factors like parental age at conception, strongly suggesting that the IVF procedure is a key factor driving the observed increase in L1 content. The more pronounced effect in females hints at potential sex-specific differences in the susceptibility or response to IVF-associated stress, possibly related to the X chromosome’s high density of L1 elements, warranting further investigation.

Beyond the quantitative difference in L1 content, we discovered qualitative differences in L1 integration patterns. We identified 11 specific genomic loci where the frequency of de novo L1 insertions was significantly different between the IVF and NC groups. Such differences are likely caused by IVF procedures rather than genetic factors. For example, we observed that the overall insertion frequency of L1Tald at the Chr15_4721515 site was higher in the naturally conceived population (insertion frequency of 40.5%) than in the IVF population (insertion frequency of 6.1%). To exclude the influence of parental genetics or other factors, we further compared the insertion status of the three pairs of IVF and naturally conceived samples from the same parents. The results showed that the insertion of L1Tal at this site was detected in the genomes of all three naturally conceived children, but not in their IVF siblings from the same parents. Similarly, at sites with higher insertion frequencies in the IVF population compared to the naturally conceived population, such as ChrX_18334634, chr2_36154109, and chr5_161008671, we observed insertions of L1 in the IVF offspring but not in the naturally conceived siblings from the same parents. Furthermore, we extended our analysis to L1 deletion events, an often overlooked aspect of L1 dynamics. We identified 14 sites with significantly different L1 deletion frequencies between the two groups, which are also likely caused by IVF procedures. For instance, at the Chr1_247687163 site, we found that L1HS was not deleted in the naturally conceived population, while 36.4% of the IVF samples had deletions at this site. Similarly, we found that the naturally conceived children did not have deletions of L1HS at this site, while their IVF siblings did in two of the three pairs of IVF and naturally conceived siblings from the same parents. It is likely that IVF procedures are responsible for these differences in L1 distribution, indicating that IVF procedures may not only promote the abnormal activation of L1 transposition but also influence the selection of L1 transposition sites during embryonic development.

Notably, a substantial portion of these differential insertions belong to the evolutionary younger and more active L1Ta and L1Ta1d subfamilies, which showed a tendency to integrate into gene-rich regions. This is particularly significant as insertions into or near genes can disrupt their function or alter their expression. Our functional analysis of the genes near these differential insertion sites revealed a remarkable convergence with the diseases reportedly more prevalent in IVF offspring. Genes associated with metabolic diseases, cardiovascular diseases, neuropsychiatric disorders, and tumors were significantly enriched in the 500-kb regions flanking these sites. Researches have been reported that L1 insertions and deletions can affect the expression of genes at or near the insertion sites, thereby promoting the occurrence and development of tumors and neuropsychiatric disorders^42–44^. For example, in this study, we found that there is a significant difference in the insertion frequency of L1 at the chr4_139726640 site on Chr4 between IVF and NC offspring populations, and this site is located in the intron region of the mastermind like transcriptional coactivator 3 (MAML3) gene. Investigation has been reported that the polymorphism of MAML3 is associated with congenital heart malformations^45^. The deletion of an L1 element can also have profound consequences, potentially remove regulatory sequences or alter the chromatin landscape. Similar to the insertion sites, the genes near these differential deletion sites were also significantly associated with metabolic, cardiovascular, neoplastic, and neuropsychiatric diseases. For instance, we found a significant difference in the deletion frequency of L1 at the chr18_52642034 site on Chr18 between IVF and NC offspring populations, and this site is located in the intron region of the DCC netrin 1 receptor (DCC) gene. Studies have been reported that the polymorphism of DCC is associated with the susceptibility to colorectal cancer and neuropsychiatric disorders such as major depressive disorder, anxiety disorder, and schizophrenia^46,47^. This indicates that IVF-associated stress may cause a dual impact on genome architecture: promoting new L1 insertions at some loci while facilitating deletions at others, both of which could contribute to genomic instability and altered gene regulation networks relevant to disease etiology.

Our study has several limitations. The sample size, while sufficient to detect significant differences, is relatively modest. The blood-derived DNA used for sequencing primarily reflects the somatic genome; thus, the L1 variations we detected are likely somatic mosaicism. It remains to be determined whether these alterations are also present in germ cells, which would have implications for trans generational inheritance. Future studies with larger cohorts, analysis of specific tissues (e.g., brain, heart), and functional validation in models are needed to establish a direct causal link between these specific L1 variants and the pathogenesis of diseases.

In conclusion, our findings demonstrate that IVF conception is associated with increased L1 content and altered patterns of L1 insertion and deletion in the offspring’s genome. The genomic regions affected by these variations are enriched for genes related to the very diseases that have been linked to IVF. This provides a novel, transposable elements centric perspective on the potential molecular underpinnings of the long-term health risks observed in the IVF population. These results underscore the importance of continuously optimizing IVF culture conditions and procedures to minimize embryonic stress. Ultimately, understanding these genomic consequences is crucial for affirming the safety of ART and guiding its future development toward generating even healthier offspring.

## Author contributions

YG, Yu L, QZ and HC designed the project. JH, ZZ, JX, YL and Yu L collected and arranged the sequencing and clinical data. JL, JH, ZZ, YG and Yu L analyzed the data. JL and ZZ wrote the manuscript. JH, QZ, HC, YG and Yu L edited the manuscript. YG and Yu L supervised the study.

## Data Availability

All data produced in the present study are available upon reasonable request to the authors

